# Derivation and validation of clinical prediction models for viral etiologies of acute diarrhea in North American children presenting for emergency care

**DOI:** 10.64898/2026.05.14.26353143

**Authors:** Paola Fonseca-Romero, Timothy Smith, Sharia M Ahmed, Anna Jones, Natalya Alekhina, Ben J. Brintz, Jennifer Dien Bard, Kimberle C. Chapin, Daniel M. Cohen, Ara Festekjian, Jami T. Jackson, Neena Kanwar, Chari D Larsen, Amy L. Leber, Rangaraj Selvarangan, Stephen Freedman, Andrew T. Pavia, Daniel T. Leung, IMPACT and APPETITE study investigators

## Abstract

**Background:** Diarrheal illness in children leads to 3.5 million care visits and 200,000 hospitalizations annually in the US. Viruses are responsible for most pediatric diarrheal cases, yet limited guidance on distinguishing viral from bacterial etiologies complicates clinical decision-making, especially regarding empiric antibiotic use.

**Methods:** We used clinical and qualitative molecular etiologic data from the Implementation of Molecular Diagnostics for Pediatric Acute Gastroenteritis (IMPACT) study to develop prediction models for viral etiology of diarrhea. We used conditional random forests to identify informative clinical and environmental predictors and evaluated model performance using logistic regression and random forests within a 5-fold cross-validation framework. We conducted external validation using the Alberta Provincial Pediatric Enteric Infection Team (APPETITE) dataset.

**Results:** Variables predictive of viral etiology included younger age, non-bloody diarrhea, winter season, and presence of vomiting. External validation showed that an AUC of 0.82 can be achieved with a parsimonious 5-variable model, yielding a sensitivity of 0.92 and specificity of 0.55

**Conclusion:** Our results suggest that in North American healthcare settings, clinical prediction models can inform decision-making by identifying children with a high probability of viral diarrhea, improving diagnostic clarity, and reducing unnecessary testing and treatment.

## Introduction

Diarrheal diseases are a major cause of illness and death among children worldwide, accounting for an estimated 1.7 billion episodes and 525,000 deaths annually in children under five^1,2^. Although the burden disproportionately affects children living in lower-resourced settings, diarrheal diseases also pose a substantial public health and economic challenge in high-income countries^3^. In the United States alone, there are approximately 178 million cases of diarrheal disease each year, resulting in 474,000 hospitalizations, 5,000 deaths, and nearly $14 billion in healthcare costs^4,5^.

Current management of pediatric diarrheal illness focuses on oral rehydration therapy, with antimicrobials beneficial only for a limited set of bacterial and protozoal etiologies^6^. The 2017 Infectious Diseases Society of America (IDSA) guidelines recommend stool testing and antimicrobial therapy only for children with severe disease, dysentery, or signs of invasive bacterial infection^6^. In the US, reported antimicrobial prescribing rates of 10-25% remain discordant with clinical recommendations, and their use is often not guided by diagnostic testing^7–9^. As a result, some children with viral etiologies of diarrhea receive unnecessary antibiotics, reflecting diagnostic uncertainty and variability in clinical practice.

In addition to challenges in determining when to use antimicrobials, the increasing use of multiplex gastrointestinal PCR panels has raised concerns about over-testing and limited clinical impact in many cases. These tests identify a broad range of organisms; however, depending on the clinical context, not all detected pathogens reflect infection, and most pathogens detected, including some bacterial pathogens, do not require antimicrobial treatment. Clinicians, therefore, also need diagnostic stewardship strategies to identify children most likely to benefit from testing ^10^.

Clinical prediction models have been developed for predicting diarrheal etiology in low- and middle-income countries (LMICs), with strong performance across diverse sites ^11,12^. However, these tools were derived using datasets from resource-limited settings and rely on clinical, nutritional, and household variables that reflect the epidemiology and care patterns of LMIC populations^11^. Notably, there is a lack of prediction models that account for the workflow and diagnostic availability in emergency departments (EDs) in high-income settings. In this study, we aimed to derive and validate clinical prediction models to identify children with a viral etiology of diarrheal illness at the time of ED presentation using data from North America-based multi-site studies of pediatric diarrhea, prioritizing variables that clinicians can readily obtain during initial assessment.

## Methods

### Study design and Setting

We conducted a secondary data analysis of pediatric patients presenting to EDs or urgent care settings with diarrheal illness in North American healthcare institutions. We followed a derivation-validation design, with model derivation and cross-validation using the Implementation of a Molecular Diagnostic for Pediatric Acute Gastroenteritis (IMPACT) dataset^13,14^. We conducted external validation using the Alberta Provincial Pediatric Enteric Infection Team (APPETITE) dataset^14^. All sites employed standardized clinical assessments and diagnostic testing for gastrointestinal pathogens. We followed the transparent reporting guidelines for multivariable prediction models for individual diagnosis (TRIPOD-AI) **(Table S1)**^15^.

The institutional review boards at participating institutions approved these studies^13,14^.

### Derivation Dataset

IMPACT was a prospective study conducted from April 2015 to September 2016 across five US pediatric EDs. Eligible patients were <18 years old and presented with symptoms of gastroenteritis (diarrhea, vomiting, abdominal pain) lasting >24 hours but <14 days. After obtaining informed consent, trained study coordinators administered a standardized questionnaire on symptoms, demographics, medical history, treatment, and epidemiologic exposure **(Table S2).** Participants submitted a stool sample at enrollment or within the next 48 hours, which was tested using a multiplex PCR platform (BioFire FilmArray GI Panel, BioMerieux, Salt Lake City, UT, USA)^16^.

### Validation Dataset

The APPETITE study, a prospective study conducted across two pediatric EDs in Alberta, Canada^14^. The study enrolled children aged <18 years presenting with acute gastroenteritis, defined as at least three episodes of vomiting or diarrhea in the preceding 24 hours and fewer than seven days of symptoms. After obtaining consent, trained staff collected standardized clinical information and obtained stool and rectal swab specimens during ED visits or through home collection kits when needed. The study team tested specimens using three laboratory methods: a five-target in-house RT-PCR viral panel, a multiplex PCR platform (Luminex xTAG Gastrointestinal Pathogen Panel, Luminex Molecular Diagnostics, ON, Canada), and standard bacterial culture^17–20^. The APPETITE study excluded children who sought medical care within the prior 14 days, unavailable for follow-up, or had conditions that prevented obtaining a rectal swab, including severe psychiatric illness and neutropenia, as well as those with diagnoses unrelated to acute gastroenteritis^14^.

### Outcome Definition

We restricted the analytic cohort to children with diarrhea who had at least one enteric pathogen detected by any of the above methods. We excluded children with no pathogen detected from the primary analysis. We defined the study outcome as viral-only diarrheal illness. Cases were classified as viral-only if testing detected at least one viral pathogen, including co-detection with more than one virus, and did not detect any bacterial or protozoal pathogen. Children with bacterial or protozoal pathogen detection, with or without viral co-detection, were classified as non-viral-only. Because detection of the *eae* gene alone (atypical EPEC) does not reliably indicate clinically significant infection in pediatric populations in high-income settings, and may reflect asymptomatic carriage, Enteropathogenic *Escherichia coli* (EPEC) was not used to define bacterial pathogen detection ^21^. Similarly, we reclassified *Clostridium difficile* detection as “not detected” in children younger than 2 years, given differences in pathogenesis and its frequent role as a colonizer in young patients^22,23^.

### Data processing

We used R version 4.4.3 to perform all data processing and analyses. We harmonized all clinical variables across the derivation and validation datasets and retained only variables available in both datasets before model development **(Table S3).** Both datasets included clinical symptoms, demographics, and vital signs available at the time of ED presentation. To reduce redundancy among predictors, we identified and removed correlated variables. We prioritized those more proximal to the patient’s clinical state (e.g., reported fever) over more distal or indirect measures (e.g., body temperature at time of presentation). We addressed missing data by imputing continuous variables with their means and categorical variables with their modes when missingness was minimal (<5% per variable). For variables with greater missingness, we introduce an “Unknown” category to preserve sample size and reduce potential bias from imputing a substantial proportion of values. For categorical predictors with more than two levels, we created dummy indicator variables for each category to enable inclusion in the modeling framework^24^.

### Clinical and environmental variables

We selected candidate clinical and environmental variables based on their clinical relevance to diarrheal disease and their availability in the ED at the time of presentation. Previous studies have shown that enteric pathogens exhibit seasonal patterns, with certain environmental conditions associated with increased detection of specific organism^25^. Prior work suggests that incorporating environmental variables may improve the clinical prediction of diarrheal disease etiology^26,27^. We defined the season according to the month of symptom onset and categorized it as winter (January-March), spring (April-June), summer (July-September), or fall (October-December).

To account for differences in seasonality in different geographic regions of the sites, we also examined the use of environmental variables in the prediction models. For the IMPACT dataset, environmental variables were assembled from publicly accessible sources: daily temperature and precipitation from the National Oceanic and Atmospheric Administration (NOAA)^28^, daily relative humidity from the Historical Weather API (Open Meteo)^29^, and air pollution data (carbon monoxide, lead, nitrogen dioxide, and ozone) from the United States Environmental Protection Agency (EPA)^30^. Missing air pollutant values (CO, Pb, NO_2_, O_3_) were addressed using a tiered imputation strategy that first applied site-specific monthly averages, followed by values from the preceding or subsequent month when monthly data were unavailable. Any remaining gaps were filled with the overall site-specific mean. For each clinical site, we extracted 14 days of environmental data per patient, including the date of symptom onset and the preceding 13 days. To account for variation in the proximity between observation stations and clinical sites, we calculated the distance from each station to the corresponding clinical site using the Haversine formula^31^. Daily weighted averages for temperature, precipitation, humidity, and air pollutants were then derived using the inverse distance weighting based on each station’s proximity to the reporting site. We handled missing weather values through mean imputation within cross-validation.

For the APPETITE dataset, we identified the nearest environmental reporting stations to the two participating sites. We obtained daily temperature data from the Government of Canada Historical Data resource^32^. We extracted hourly carbon monoxide concentrations from the Government of Canada National Air Pollution Surveillance (NAPS) system using site-specific geographic coordinates and then converted to daily means ^33,34^.

### Statistical modeling and assessment

We fit conditional random forest models (cforest) and measured predictor importance using area under the receiver operating characteristic curve (AUC)-based conditional permutation importance. We ranked variables by importance and included the top predictors in logistic regression and random forest classification models. Within the five-fold cross-validation framework, we randomly partitioned the data into five subsets: four for model training and one for testing. We repeated this process so that each fold served as the test set exactly once.

Within each training fold, we identified the highest-ranking predictors on each training fold and evaluated subsets containing the top two through 10 predictors.

We applied this approach to two categories of models: (1) clinical predictor-only models using demographic and symptom-based variables, and (2) “clinical + environmental” models incorporating 14-day aggregate temperature, humidity, and pollutant exposures. For both outcomes, we fit regression models using the selected predictors and random forest classifiers with 1,000 trees. We evaluated model performance using 5-fold cross-validation and calculated the mean AUC across folds. The AUC measures the model’s ability to distinguish between children with viral-only and non-viral etiologies, where value of 0.5 indicate no discriminative ability and a value of 1.0 indicates perfect discrimination. To classify individual patients, we applied a probability threshold, defined as the cut-off above which a child was classified as having a viral-only etiology. We evaluated thresholds of 0.49, selected during cross-validation, and explore other thresholds to assess the tradeoff between sensitivity and specificity. We also calculated positive predictive value (PPV) and negative predictive value (NPV) to assess classification performance.

### Model Validation

Based on the 5-fold cross-validation results, we refit the best-performing models for each analysis on the full IMPACT dataset and evaluated external performance using the APPETITE dataset. In addition to predictive performance, we report odds ratios for each predictor from the logistic regression models to better understand the direction and magnitude of the predictor’s effect on the final probability for viral etiology.

We assessed model calibration using calibration curves to evaluate agreement between predicted and observed probabilities of viral-only etiology. We evaluated clinical utility with decision curves analysis using viral -only etiology as a positive class ^35–37^.

## Results

### Study Population and Pathogen Distribution

Of 1,157 children enrolled in the IMPACT study, 1020 met criteria for acute diarrhea **(Figure S1).** Multiplex PCR results were -11available for 868 children **(Table S4),** among whom 570 (66%) had at least one pathogen detected **(Table S5).** Of these, 337 (59%) had viral-only detection and 233 (41%) had other etiologies, including bacterial, protozoal, and mixed detections **(Table S6).** Of 1,090 children enrolled in the APPETITE study with diarrhea, 1,088 (99.8%) underwent testing with Luminex xTAG and RT-PCR viral assays **(Table S7).** Among those tested, 830 had ≥1 pathogen detected, of whom 745 (90%) had viral-only detection. **(Table S8).**

When stratifying by age, we found that viral pathogens were most prevalent among younger children, particularly those <24 months of age; non-viral etiologies occurred more often in older children (**Table 1).** In univariate analysis, vomiting was more prevalent among children with viral etiology in both datasets, while fever and bloody diarrhea were consistently more frequent in non-viral infections. Seasonal patterns also differed: non-viral infections occurred more commonly during the summer months in both datasets. Additionally, among environmental variables, higher average temperature was associated with non-viral infections in IMPACT, while average carbon monoxide levels were slightly higher among viral episodes.

**Table 1.** Baseline characteristics of participants stratified by viral and non-viral etiology in the IMPACT and APPETITE datasets.

| Variables | IMPACT<br>Viral<br>(n = 337)<sup>1</sup> | IMPACT<br>Non-viral<br>(n = 233)<sup>1</sup> | APPETITE<br>Viral<br>(n = 745)<sup>1</sup> | APPETITE<br>Non-viral<br>(n = 85)<sup>1</sup> |
| --- | --- | --- | --- | --- |
| **Age category** |  |  |  |  |
| < 6 months | 46 (14%) | 9 (3.9%) | 51 (6.8%) | 1 (1.2%) |
| 6-23 months | 147 (44%) | 54 (23%) | 414 (56%) | 25 (29%) |
| 2-4 years | 71 (21%) | 69 (30%) | 216 (29%) | 30 (35%) |
| 5-11 years | 54 (16%) | 69 (30%) | 58 (7.8%) | 26 (31%) |
| 12-17 years | 19 (5.6%) | 32 (14%) | 6 (0.8%) | 3 (3.5%) |
| **Vomiting/Nausea** | 288 (85%) | 164 (70%) | 651 (87%) | 55 (65%) |
| **Fever** | 152 (45%) | 153 (66%) | 410 (55%) | 58 (68%) |
| **Duration of diarrhea**<br>**(days)** | 3.00<br>(1.00, 5.0) | 3.00<br>(2.00, 4.00) | 3.00<br>(1.00, 4.00) | 3.00<br>(2.00, 4.00) |
| **Bloody diarrhea** | 14 (4.2%) | 55 (24%) | 27 (3.6%) | 30 (35%) |
| **Number of vomiting**<br>**in the last 24 hours** |  |  |  |  |
| High (≥ 3 episodes) | 211 (63%) | 142 (61%) | 484 (65%) | 28 (33%) |
| Low (< 3 episodes) | 126 (37%) | 91 (39%) | 261 (35%) | 57 (67%) |
| **School/daycare**<br>**exposure** | 173 (51%) | 143 (61%) | 274 (37%) | 23 (27%) |
| **Season** |  |  |  |  |
| Winter | 115 (34%) | 43 (18%) | 204 (27%) | 19 (22%) |
| Spring | 57 (17%) | 37 (16%) | 296 (40%) | 24 (28%) |
| Summer | 64 (19%) | 83 (36%) | 117 (16%) | 27 (32%) |
| Fall | 101 (30%) | 70 (30%) | 128 (17%) | 15 (18%) |
| **Household members**<br>**aged 5-15 years** | 222 (66%) | 178 (76%) | 454 (61%) | 62 (73%) |
| Prior care-seeking for<br>same illness | 115 (34%) | 69 (30%) | 173 (23%) | 24 (28%) |
| **Environmental**<br>**Carbon monoxide**<br>**(14-day avg)<sup>2</sup>** | 0.64<br>(0.41, 0.76) | 0.48<br>(0.37, 0.66) | 0.20<br>(0.16, 0.24) | 0.20<br>(0.15, 0.23) |
| **Environmental**<br>**Temperature (14-day**<br>**avg)<sup>2</sup>** | 52 (42, 67) | 63 (49, 74) | 44<br>(30, 56) | 47<br>(30, 61) |

<sup>1</sup>n (%) for categorical variables; Median (Q1, Q3) for continuous variables.
<sup>2</sup>Environmental variables represent ambient measurements aggregated over the 14 days prior to presentation. Environmental carbon monoxide (CO) is reported in parts per million (ppm), and environmental temperature is reported in degrees Fahrenheit (°F).

### Model Development and Internal Performance

In our model derived solely from clinical variables, age emerged as the strongest predictor of viral etiology, and removing age reduced the AUC by 0.0517 **(Table 2)**. The absence of bloody diarrhea and fever, the presence of vomiting/nausea, and non-summer season also ranked among the top predictors. Additional variables with lesser but notable importance included the duration of diarrhea symptoms and household members aged 5-15 years. In contrast, prior ED care, the number of vomiting episodes in 24 hours, and daycare attendance contributed minimally to model performance. In the clinical plus environmental model, environmental variables contributed meaningfully to prediction, with mean temperature (14-day average) and carbon monoxide ranking fourth (0.0088) and sixth (0.0057), respectively, in importance **(Table 2),** while season ranked ninth. Partial dependency plots demonstrated that higher carbon monoxide levels were associated with an increased predicted probability of viral etiology, whereas higher temperatures were associated with a decreased probability of viral etiology **(Figure S2)**

**Table 2:** Top 10 predictors for viral-only etiology based on AUC-based conditional permutation importance in the clinical-only model and clinical + environmental model.

| Rank | Clinical-only Model | Importance | Clinical + Environmental Model | Importance |
| --- | --- | --- | --- | --- |
| 1 | Age (months) | 0.0517 | Age (months) | 0.0362 |
| 2 | Bloody diarrhea | 0.0272 | Bloody diarrhea | 0.0236 |
| 3 | Fever | 0.0185 | Fever | 0.0150 |
| 4 | Summer season | 0.0137 | Mean temperature (14-day avg) | 0.0088 |
| 5 | Vomiting or nausea | 0.0153 | Vomiting or nausea | 0.0087 |
| 6 | Duration of diarrhea (days) | 0.0026 | Carbon monoxide (14-day avg) | 0.0057 |
| 7 | Household members aged 5-15 years | 0.0030 | Household members aged 5-15 years | 0.0027 |
| 8 | Prior healthcare visit in the ED | 0.0015 | Duration of diarrhea (days) | 0.0013 |
| 9 | Frequency of vomiting in 24 hours | 0.0025 | Summer season | 0.0013 |
| 10 | Day care attendance | 0.0001 | Prior healthcare visit in the ED | 0.0008 |

Importance scores represent AUC-based conditional permutation importance, where larger values
indicate a greater reduction in model performance when the variable is permuted.

Using 5-fold cross-validation, we evaluated logistic regression and random forest models for predicting viral etiology. We observed mean AUCs ranging from 0.71 for models with 1 predictor to 0.76 for models with 5 or more predictors **(Figure 1).** In clinical-only models, mean AUC increased from approximately 0.72 with two predictors to 0.77-0.78 with eight predictors for both logistic regression and random forest approaches **(Figure 1A, Figure S3).**

**Figure 1:**
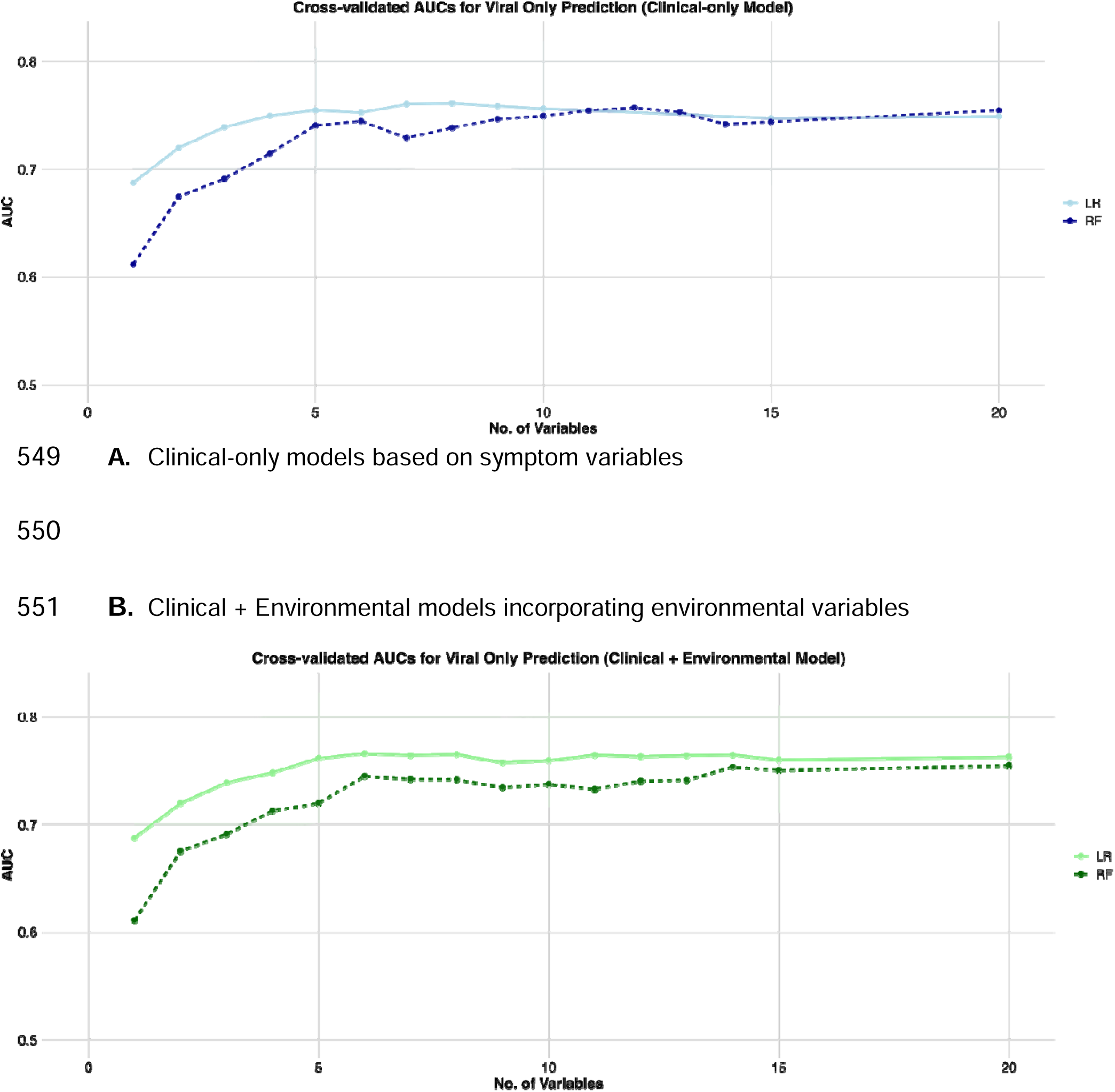
Cross-validated discrimination of clinical-only and clinical + environmental models. 5-fold cross-validation of random forest classification and logistic regression model according to the number of predictors used in each model (AUC vs number of variables). The solid line represents logistic regression (LR) models, and the dashed line represents random forest (FR) models. Shaded regions represent variability across folds (mean ± SD).

Logistic regression models consistently showed slightly higher discrimination than random forest across predictor subsets. **(Figure 1B, Figure S4).** Incorporating environmental variables further improved performance. In these models, mean AUC increased from 0.75 with four predictors to 0.79 with seven predictors, and logistic regression again outperformed random forests.

In the derivation models, younger age was strongly associated with viral etiology (OR 0.87 per month, 95% CI 0.82-0.92, p<0.001), whereas bloody diarrhea (OR 0.18, 95% CI 0.09-0.34) and fever (OR 0.35, 95% CI 0.24-0.53) were associated with lower odds of viral detection **(Table 3).** In contrast, vomiting/nausea was associated with higher odds of viral etiology (OR 3.04, 95% CI 1.77-05.26) while summer season was associated with reduced odds (OR 0.42, 95% CI 0.27-0.66).

**Table 3:** The odds ratios, 95% confidence interval, and p-value from the logistic regression model for clinical-only model and clinical + environmental model for viral-only outcome.

| Variable | OR (95% CI)<br>(Clinical-only model) | p-value | OR (95% CI)<br>(Clinical + Environmental) | p-value |
| --- | --- | --- | --- | --- |
| Age (months) | **0.87 (0.82-0.91)** | < 0.001 | **0.86 (0.82-0.91)** | < 0.001 |
| Bloody diarrhea | **0.18 (0.09-0.34)** | < 0.001 | **0.17 (0.08-0.33)** | < 0.001 |
| Fever | **0.35 (0.24-0.53)** | < 0.001 | **0.37 (0.24-0.55)** | < 0.001 |
| Summer season | **0.42 (0.27-0.66)** | < 0.001 | 0.74 (0.41-1.34) | 0.3 |
| Vomiting or nausea | **3.04 (1.77-5.26)** | < 0.001 | **2.36 (1.45-3.86)** | < 0.001 |
| Duration of diarrhea (days) | 0.98 (0.90-1.06) | 0.6 | 0.96 (0.89-1.03) | 0.2 |
| Household members aged 5-15 years | 1.68 (0.80-3.50) | 0.2 | 1.61 (0.76-3.37) | 0.2 |
| Prior healthcare visit in the ED | **1.58 (1.02-2.46)** | 0.07 | 1.52 (0.98-2.37) | 0.062 |
| Frequency of vomiting in 24 hours | 0.66 (0.42-1.04) | 0.076 | — | — |
| Day care attendance | 0.95 (0.61-1.49) | 0.8 | — | — |
| Carbon monoxide (14-day avg) | — | — | 2.13 (0.77-6.00) | 0.15 |
| Temperature (14-day avg) | — | — | **0.98 (0.96-1.00)** | 0.034 |

<sup>1</sup>Reference category for binary variables in “No”; for categorical variables, the first level (e.g., winter for season) was used as the reference.
<sup>2</sup>Abbreviations: CI = Confidence Interval, OR = Odds Ratio

In our cross-validation model, we observed a consistent direction of effect for the top predictors. Mean temperature showed a modest inverse association (OR 0.98, 95% CI 0.96-1.00), while carbon monoxide and other variables were not statistically significant **(Table 3).** Overall, these findings demonstrate stable and consistent predictor effects across cohorts.

### External Validation Performance

In external validation using the APPETITE dataset, LR and RF clinical-only models achieved higher performance than during internal cross-validation, with 5-predictor models consistently demonstrating the strongest discriminative performance across analyses. Using clinical-only predictors **(Figure 2A),** LR achieved the highest performance using the top five predictors (AUC = 0.81). In comparison, models with three and ten predictors showed slightly lower discrimination (AUC = 0.76 and 0.79, respectively) **(Table S9).** At the prespecified probability threshold of 0.49, the five-predictor LR model achieved a sensitivity of 0.91 and specificity of 0.55. RF models demonstrated comparable performance, with AUCs ranging from 0.76 to 0.79. Overall, models using five predictors achieved the best discrimination, and increasing model complexity beyond this threshold did not substantially improve performance in the external dataset.

**Figure 2:**
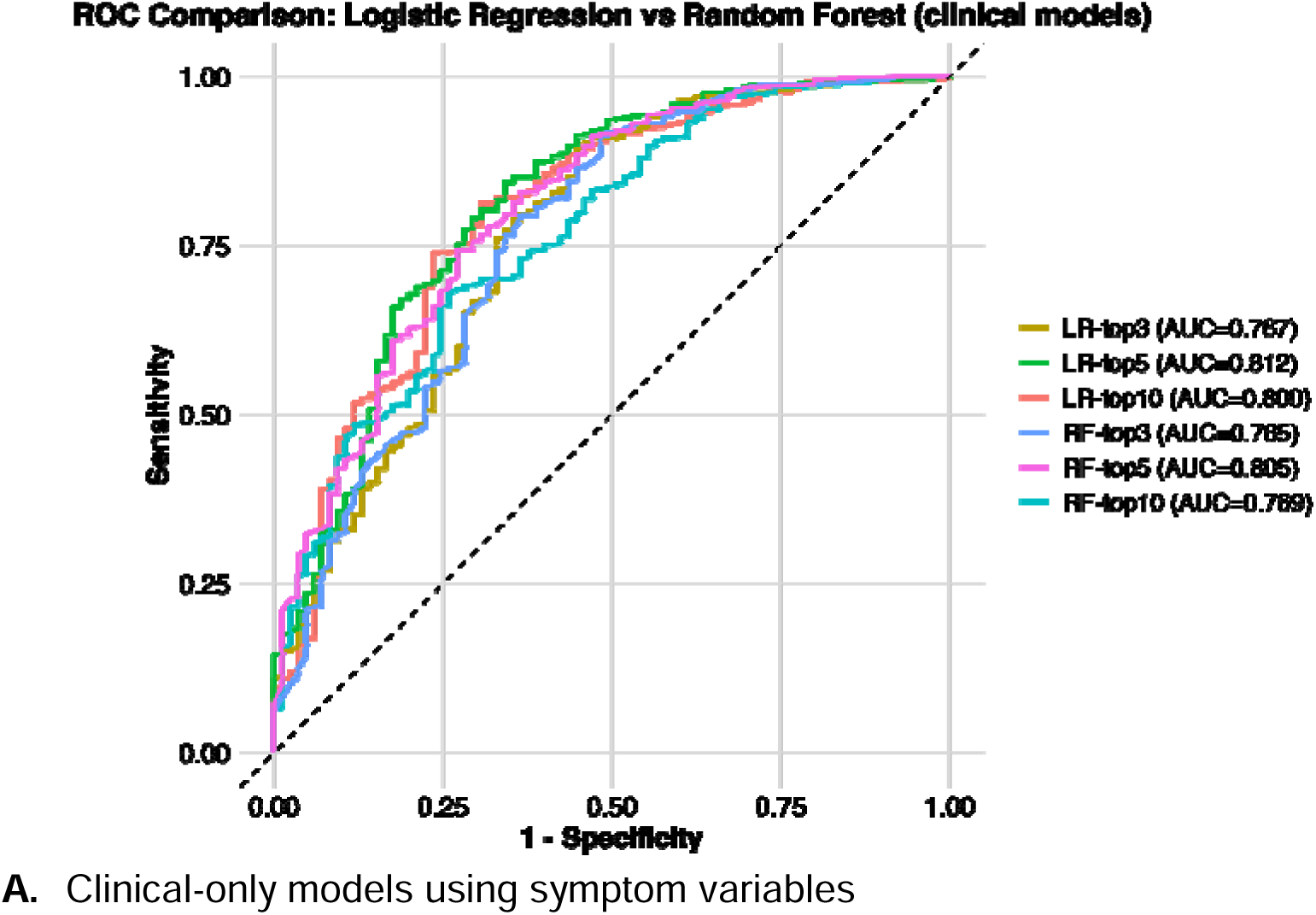
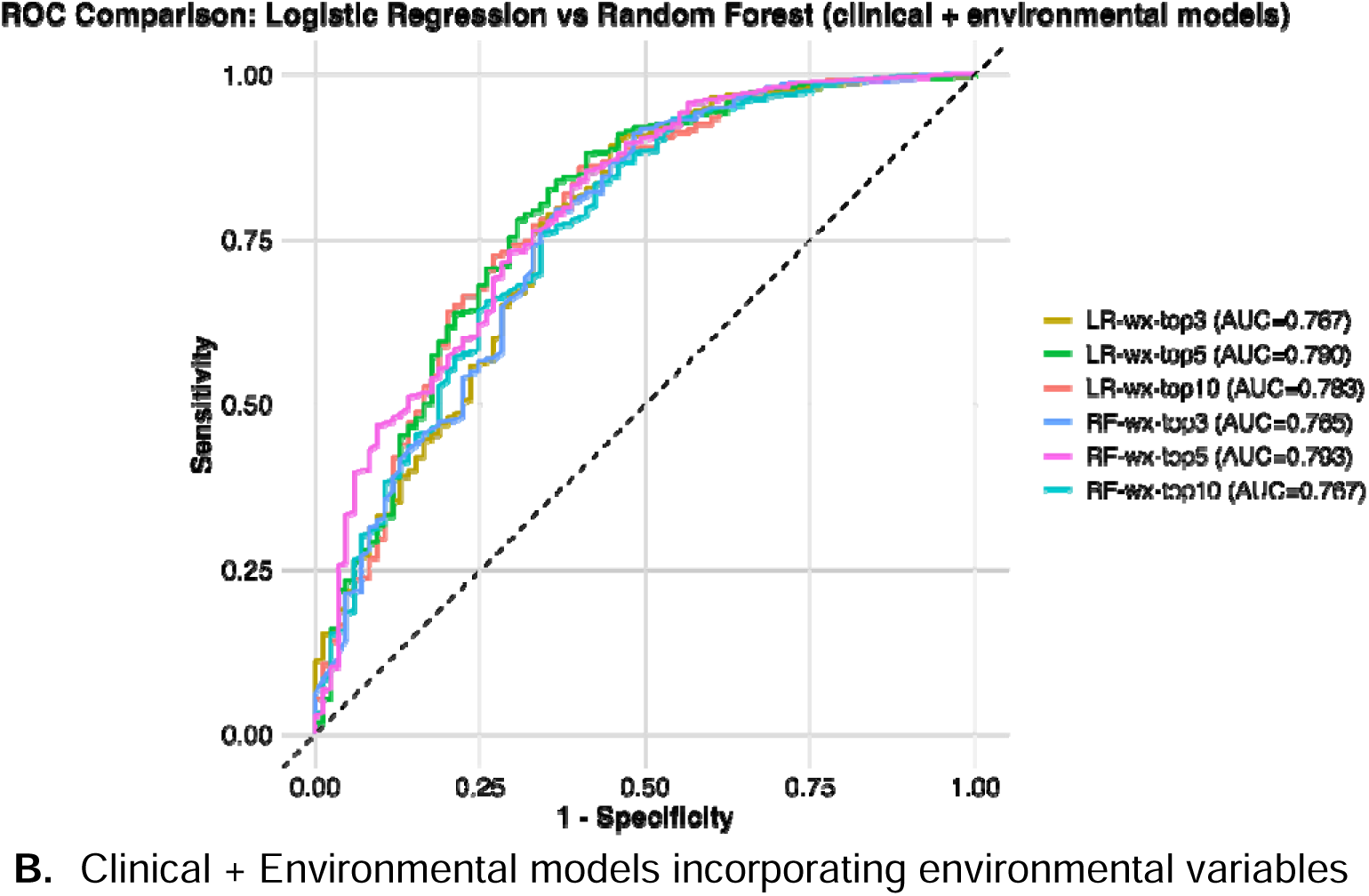
External validation receiver operating characteristic (ROC) curves for logistic regression (LR) and random forest (RF) models using the APPETITE dataset. Colors represent models using different numbers of top-ranked predictors (3, 5, and 10). The dashed diagonal line indicates no discriminative ability.

### Calibration and Clinical Utility

Calibration assessment in the external validation demonstrated differences between predicted and observed risk across models **(Figure 3).** For the LR model using clinical-only, calibration intercepts were consistently positive (range: 1.63-1.94), indicating systemic underprediction of viral etiology in the external dataset. This finding was consistent with the calibration plots, which showed that observed probabilities were generally higher than predicted across risk strata.

**Figure 3:**
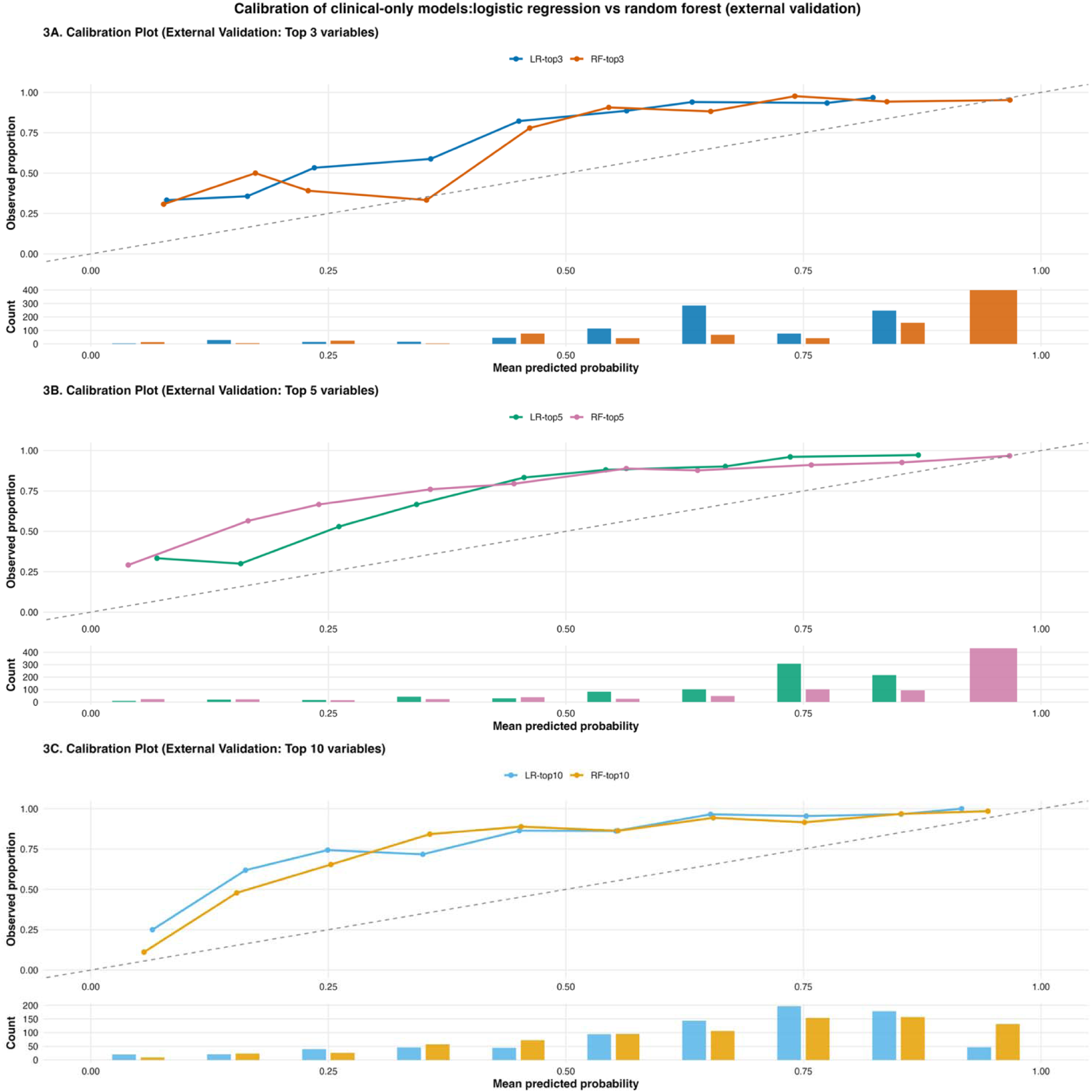
External validation calibration plots for clinical-only prediction models using 3, 5, and 10 predictors (APPETITE dataset).

Calibration plots consistently showed higher observed than predicted probabilities across risk strata. Calibration slopes remain close to 1 (1.00-1.27), indicating reasonable agreement between predicted and observed risk gradients, although models with fewer variables demonstrated slight overfitting.

In contrast, RF models also demonstrated positive calibration intercepts (range: 1.34-1.80), indicating underprediction similar to LR, but showed substantially lower calibration slopes (range 0.57-0.92). The observed underprediction across all models is likely attributable to differences in outcome prevalence between the derivation and validation datasets, with a higher proportion of viral cases in the external validation cohort (APPETITE: ∼89% viral vs IMPACT: ∼59% viral).

To assess the clinical utility of our models, we performed decision curve analysis (**Figure 4)** to determine whether model-guided diagnostic testing strategies could improve clinical decision-making compared with testing all patients or no testing. In this analysis, we defined viral-only etiology as the positive class. A positive model prediction indicated a high probability of viral diarrhea, which could support deferring diagnostic testing in low-risk patients. The “test all” strategy assumed diagnostic testing for all patients, whereas the “test none” strategy assumed no diagnostic testing. Decision curve analysis demonstrated that all models provided greater net benefit than the test-none strategy across a broad range of threshold probabilities. Compared with the test-all strategy, models demonstrated greater clinical utility at moderate threshold probabilities (approximately 0.4-0.8). LR models incorporating 5 and 10 predictors demonstrated the most stable net benefit across threshold probabilities.

**Figure 4:**
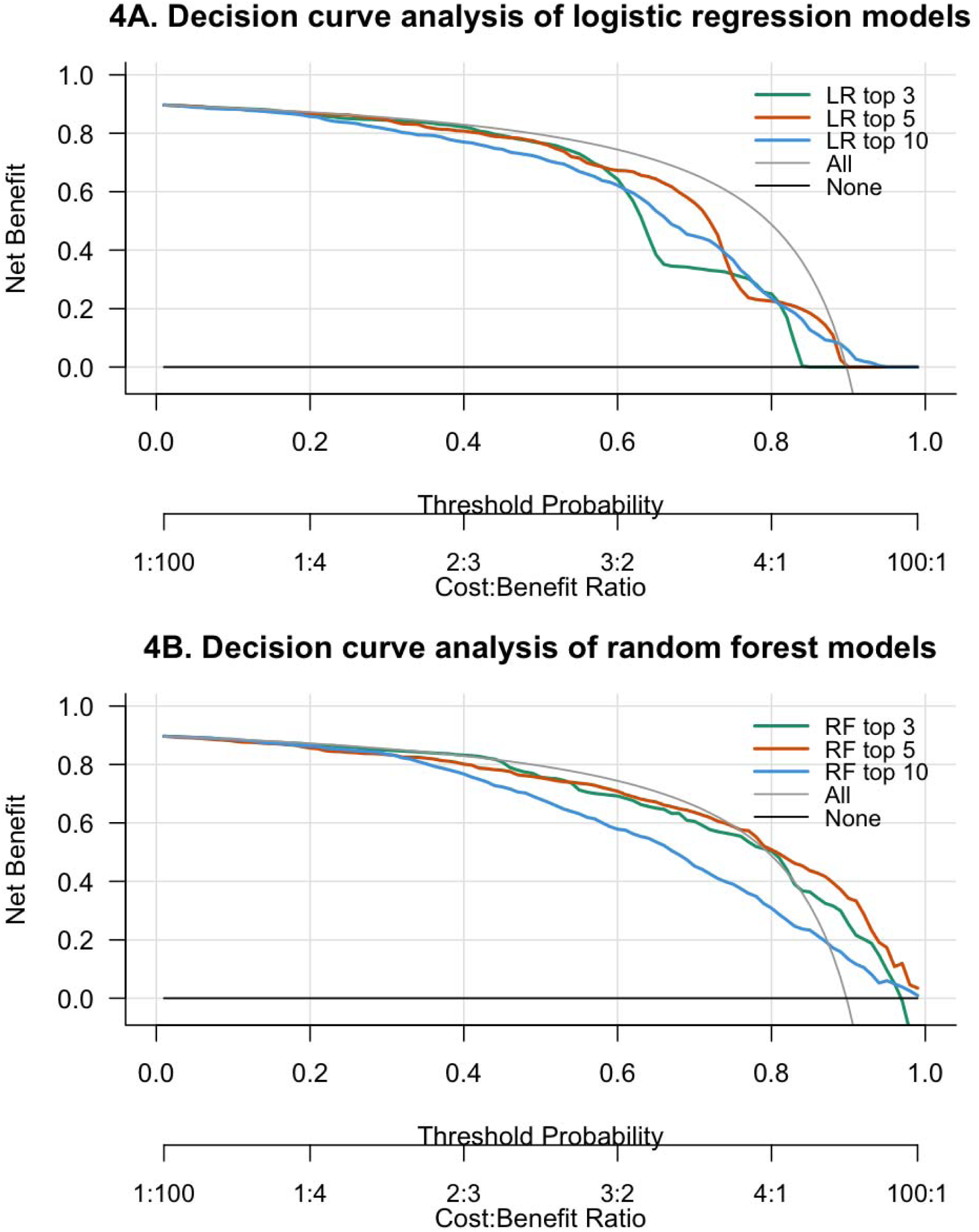
Decision curve analysis of clinical-only logistic regression and random forest models for guiding diagnostic testing strategies in children with diarrhea **Footnote:** Decision curves display net benefit across a range of threshold probabilities with the corresponding cost; benefit shown on the secondary x-axis. The threshold probability is the predicted probability of viral-only etiology at which diagnostic testing would be deferred. The reference strategies are testing all patients (“Test all”) and testing no patients (“Test none”).

## Discussion

In this study, we developed and externally validated clinical prediction models to distinguish viral from non-viral etiology of diarrhea among children presenting to EDs, using readily available clinical and environmental variables. We identified younger age, non-bloody diarrhea, absence of fever, vomiting, and shorter diarrhea duration as the strongest predictors of viral infection. A parsimonious model using these five clinical predictors accurately predicted viral etiology, achieving an externally validated AUC of 0.82 in an independent dataset that employed a different multiplex diagnostic testing approach. These findings demonstrate that a small number of predictors can achieve strong discrimination, supporting the development of practical tools to guide diagnostic testing in North American settings.

Enhancing clinicians’ ability to predict viral infection reliably should further reduce the inappropriate use of antibiotics in children with acute diarrhea. In addition, it has the potential to minimize unnecessary diagnostic testing and reduce the risk to patients from the adverse effects of unnecessary antibiotics. In our analysis, we prioritized diagnostic specificity for a “viral only” etiology to minimize the risk of withholding diagnostic testing from children with bacterial infections. Our top predictors align with known factors associated with viral etiology of pediatric diarrheal disease, including young age^38,39^, and non-bloody diarrhea or fever ^11^. Vomiting and seasonal clustering, particularly during winter months when rotavirus and norovirus circulate, are also associated with viral etiology^38,39^.

Environmental variables identified as important predictors in cross-validation outperformed a crude seasonality variable and reflect known associations between weather conditions and enteric pathogen transmission^26,27,40^. Lower ambient temperature and higher carbon monoxide levels were the top predictors of viral etiology. However, these findings likely capture seasonal transmission patterns and behavioral factors, such as increased indoor crowding, rather than direct effects on viral pathogenicity. Although incorporating environmental variables did not improve external validation performance, our results suggest that future etiologic prediction models for infectious diseases should still consider environmental factors, which may generalize across geographic regions more effectively than seasonal variables.

The clinical variables in our clinical prediction model could be entered into an app or captured from the history using large language models, generating a probability of viral infection. A potential implementation of this model could involve integration into the electronic health record as a “pop up” tool for patients with a chief complaint of diarrhea or when ordering stool testing. The probabilities could help clinicians decide whether additional molecular testing would likely change management and whether empiric antimicrobials were warranted.

This study has several limitations. Diagnostic testing methods differed between the derivation and validation cohorts, which may have influenced pathogen detection rates and contributed to outcome misclassification. In addition, multiplex PCR assays can detect low-level viral shedding or asymptomatic carriage, particularly for rotavirus in recently vaccinated children, which may not always represent the cause of illness. Future studies should prospectively validate these models in diverse clinical settings, assess their impact on diagnostic testing and antibiotic use.

In summary, we proposed a five-predictor model to identify children with a viral-only etiology of their diarrhea. Usage of this model could improve antibiotic and diagnostic stewardship in North American settings. This model could be integrated into an application-based tool to provide real-time decision-making guidance for diagnostic testing in children with acute diarrhea.

## Acknowledgements

We thank all the support staff who participated in this project. The IMPACT study investigators include: Andrew T. Pavia, Daniel M. Cohen, Amy L. Leber, Judy A. Daly, Jami T. Jackson, Rangaraj Selvarangan, Neena Kanwar, Jeffrey M. Bender, Jennifer Dien Bard, Ara Festekjian, Susan Duffy, Chari D. Larsen, Kristen M. Holmberg, Tyler Bardsley, Benjamin Haaland, Kevin M. Bourzac, Christopher Stockmann, Kimberle C. Chapin, and Daniel T. Leung. The APPETITE study investigators include: Bonita Lee, Marie Louie, Xiao-Li Pang, Samina Ali, Andy Chuck, Linda Chui, Gillian Currie, James Dickinson, Steven Drews, Mohamed Eltorki, Tim Graham, Xi Jiang, David Johnson, James Kellner, Martin Lavoie, Judy MacDonald, Shannon MacDonald, Lawrence Svenson, James Talbot, Phillip Tarr, Raymond Tellier, Otto Vanderkooi.

## Data availability

A limited de-identified dataset and analytic code are available at [TBD].

