## Supplementary Materials for "Derivation and validation of clinical prediction models for viral etiologies of acute diarrhea in North American children presenting for emergency care"

**Supplementary Table 1:** Checklist TRIPOD+AI

| **Section/Topic Item Development Checklist item**  **/ evaluation**^1^ | | | | | | | | **Reported on page**  **7**  **1**  **3**  **4**  **5**  **5**  **3**  **5**  **5**  **5**  **N/A**  **5**  **6**  **N/A**  **7**  **7**  **N/A** |
| --- | --- | --- | --- | --- | --- | --- | --- | --- |
| **TITLE** | | | | | | | |  |
| *Title* | | 1 | | D;E | | Identify the study as developing or evaluating the performance of a multivariable prediction model, the target population, and the outcome to be predicted | |  |
| **ABSTRACT** | | | | | | | | |
| *Abstract* | | 2 | | D;E | | See TRIPOD+AI for Abstracts checklist | |  |
| **INTRODUCTION** | | | | | | | | |
| *Background* | | 3a | | D;E | | Explain the healthcare context (including whether diagnostic or prognostic) and rationale for developing or evaluating the prediction model, including references to existing models | |  |
|  |  | 3b | | D;E | | Describe the target population and the intended purpose of the prediction model in the context of the care pathway, including its intended users (e.g., healthcare professionals, patients, public) | |  |
|  |  | 3c | | D;E | | Describe any known health inequalities between sociodemographic groups | |  |
| *Objectives* | | 4 | | D;E | | Specify the study objectives, including whether the study describes the development or validation of a prediction model (or both) | |  |
| **METHODS** | | | | | | | | |
| *Data* | | 5a | | D;E | | Describe the sources of data separately for the development and evaluation datasets (e.g., randomised trial, cohort, routine care or registry data), the rationale for using these data, and representativeness of the data | |  |
|  |  | 5b | | D;E | | Specify the dates of the collected participant data, including start and end of participant accrual; and, if applicable, end of follow-up | |  |
| *Participants* | | 6a | | D;E | | Specify key elements of the study setting (e.g., primary care, secondary care, general population)  including the number and location of centres | |  |
|  |  | 6b | | D;E | | Describe the eligibility criteria for study participants | |  |
|  |  | 6c | | D;E | | Give details of any treatments received, and how they were handled during model development or evaluation, if relevant | |  |
| *Data preparation* | | 7 | | D;E | | Describe any data pre-processing and quality checking, including whether this was similar across  relevant sociodemographic groups | |  |
| *Outcome* | | 8a | | D;E | | Clearly define the outcome that is being predicted and the time horizon, including how and when assessed, the rationale for choosing this outcome, and whether the method of outcome assessment is  consistent across sociodemographic groups | |  |
|  |  | 8b | | D;E | | If outcome assessment requires subjective interpretation, describe the qualifications and demographic characteristics of the outcome assessors | |  |
|  |  | 8c | | D;E | | Report any actions to blind assessment of the outcome to be predicted | |  |
| *Predictors* | | 9a | | D | | Describe the choice of initial predictors (e.g., literature, previous models, all available predictors) and  any pre-selection of predictors before model building | |  |
|  |  | 9b | | D;E | | Clearly define all predictors, including how and when they were measured (and any actions to blind assessment of predictors for the outcome and other predictors) | |  |
|  |  | 9c | | D;E | | If predictor measurement requires subjective interpretation, describe the qualifications and demographic characteristics of the predictor assessors  **N/A** | |  |
| *Sample size* | | 10 | | D;E | | Explain how the study size was arrived at (separately for development and evaluation), and justify that  the study size was sufficient to answer the research question. Include details of any sample size calculation  **6** | |  |
| *Missing data* | | 11 | | D;E | | Describe how missing data were handled. Provide reasons for omitting any data  **7** | |  |
| *Analytical methods* | | 12a | | D | | Describe how the data were used (e.g., for development and evaluation of model performance) in the analysis, including whether the data were partitioned, considering any sample size requirements  **5** | |  |
|  |  | 12b | | D | | Depending on the type of model, describe how predictors were handled in the analyses (functional form,  rescaling, transformation, or any standardisation).  **7-8** | |  |
|  |  | 12c | | D | | Specify the type of model, rationale^2^, all model-building steps, including any hyperparameter tuning,  **9-10**  and method for internal validation | |  |
|  |  | 12d | | D;E | | Describe if and how any heterogeneity in estimates of model parameter values and model performance was handled and quantified across clusters (e.g., hospitals, countries). See TRIPOD-Cluster for  **N/A**  additional considerations^3^ | |  |
|  |  | 12e | | D;E | | Specify all measures and plots used (and their rationale) to evaluate model performance (e.g., discrimination, calibration, clinical utility) and, if relevant, to compare multiple models  **9** | |  |
|  |  | 12f | | E | | Describe any model updating (e.g., recalibration) arising from the model evaluation, either overall or for particular sociodemographic groups or settings  **10** | |  |
|  |  | 12g | | E | | For model evaluation, describe how the model predictions were calculated (e.g., formula, code, object, application programming interface)  **9** | |  |
| *Class imbalance* | | 13 | | D;E | | If class imbalance methods were used, state why and how this was done, and any subsequent methods to  **N/A**  recalibrate the model or the model predictions | |  |
| *Fairness* | | 14 | | D;E | | Describe any approaches that were used to address model fairness and their rationale  **N/A** | |  |
| *Model output* | | 15 | | D | | Specify the output of the prediction model (e.g., probabilities, classification). Provide details and  **9**  rationale for any classification and how the thresholds were identified | |  |
| *Training versus*  *evaluation* | | 16 | | D;E | | Identify any differences between the development and evaluation data in healthcare setting, eligibility  **5-6**  criteria, outcome, and predictors | |  |
| *Ethical approval* | | 17 | | D;E | | Name the institutional research board or ethics committee that approved the study and describe the participant-informed consent or the ethics committee waiver of informed consent  **5** | |  |
| **OPEN SCIENCE** | | | | | | | | |
| *Funding* | 18a | | D;E | | Give the source of funding and the role of the funders for the present study  **26** | |  | |
| *Conflicts of interest* | 18b | | D;E | | Declare any conflicts of interest and financial disclosures for all authors  **26** | |  | |
| *Protocol* | 18c | | D;E | | Indicate where the study protocol can be accessed or state that a protocol was not prepared  **5** | |  | |
| *Registration* | 18d | | D;E | | Provide registration information for the study, including register name and registration number, or state  **5**  that the study was not registered | |  | |
| *Data sharing* | 18e | | D;E | | Provide details of the availability of the study data  **26** | |  | |
| *Code sharing* | 18f | | D;E | | Provide details of the availability of the analytical code^4^  **26** | |  | |
| **PATIENT & PUBLIC INVOLVEMENT** | | | | | | | | |
| *Patient & Public Involvement* | 19 | | D;E | | Provide details of any patient and public involvement during the design, conduct, reporting, interpretation, or dissemination of the study or state no involvement.  **N/A** | |  | |
| **RESULTS** | | | | | | | | |
| *Participants* | 20a | | D;E | | Describe the flow of participants through the study, including the number of participants with and without the outcome and, if applicable, a summary of the follow-up time. A diagram may be helpful.  **10-11** | |  | |
|  | 20b | | D;E | | Report the characteristics overall and, where applicable, for each data source or setting, including the key dates, key predictors (including demographics), treatments received, sample size, number of outcome events, follow-up time, and amount of missing data. A table may be helpful. Report any  **10-11**  differences across key demographic groups. | |  | |
|  | 20c | | E | | For model evaluation, show a comparison with the development data of the distribution of important predictors (demographics, predictors, and outcome).  **11** | |  | |
| *Model development* | 21 | | D;E | | Specify the number of participants and outcome events in each analysis (e.g., for model development, hyperparameter tuning, model evaluation)  **11** | |  | |
| *Model specification* | 22 | | D | | Provide details of the full prediction model (e.g., formula, code, object, application programming interface) to allow predictions in new individuals and to enable third-party evaluation and implementation, including any restrictions to access or re-use (e.g., freely available, proprietary)^5^  **6** | |  | |
| *Model performance* | 23a | | D;E | | Report model performance estimates with confidence intervals, including for any key subgroups (e.g., sociodemographic). Consider plots to aid presentation.  **18** | |  | |
|  | 23b | | D;E | | If examined, report results of any heterogeneity in model performance across clusters. See TRIPOD  **N/A**  Cluster for additional details^3^. | |  | |
| *Model updating* | 24 | | E | | Report the results from any model updating, including the updated model and subsequent performance  **N/A** | |  | |
| **DISCUSSION** | | | | | | | | |
| *Interpretation* | 25 | | D;E | | Give an overall interpretation of the main results, including issues of fairness in the context of the  **12-18**  objectives and previous studies | |  | |
| *Limitations* | 26 | | D;E | | Discuss any limitations of the study (such as a non-representative sample, sample size, overfitting, missing data) and their effects on any biases, statistical uncertainty, and generalizability  **25** | |  | |
| *Usability of the model in the context of current care* | 27a | | D | | Describe how poor quality or unavailable input data (e.g., predictor values) should be assessed and handled when implementing the prediction model  **24-25** | |  | |
|  | 27b | | D | | Specify whether users will be required to interact in the handling of the input data or use of the model,  **25**  and what level of expertise is required of users | |  | |
|  | 27c | | D;E | | Discuss any next steps for future research, with a specific view to applicability and generalizability of  **25**  the model | |  | |

From: Collins GS, Moons KGM, Dhiman P, et al. BMJ 2024;385:e078378. doi:10.1136/bmj-2023-078378

______________________________________________

^1^ D=items relevant only to the development of a prediction model; E=items relating solely to the evaluation of a prediction model; D;E=items applicable to both the development and evaluation of a prediction model

^2^ Separately for all model building approaches.

^3^ TRIPOD-Cluster is a checklist of reporting recommendations for studies developing or validating models that explicitly account for clustering or explore heterogeneity in model performance (eg, at different hospitals or centres). Debray et al, BMJ 2023; 380: e071018 [DOI: 10.1136/bmj-2022-071018]

^4^ This relates to the analysis code, for example, any data cleaning, feature engineering, model building, evaluation.

^5^ This relates to the code to implement the model to get estimates of risk for a new individual.

**Supplementary Table 2:** Variable available to collect in emergency department (ED)

| Variables available in ED | | | |
| --- | --- | --- | --- |
| Subject Age | Loss of appetite | International travel in past month | Immunosuppressive condition |
| Sex | Fatigue/weakness | Chills | Duration of diarrhea at presentation |
| Race | Body aches | Vomiting/nausea | Insurance |
| Ethnicity | Difficulty breathing | Contact with animal/pet in last month | Rehydration |
| Season | Bloody diarrhea | Drank well water in past month | Excessive crying/fussiness |
| Abdominal pain | Watery diarrhea | Number of people in subject's household <5 years old | Out of state travel in past month |
| Duration of vomiting at presentation | Mucousy diarrhea | Number of people in subject's household 5-17 years old | Diabetes |
| Number of stools in 24 hours prior to presentation | Duration of diarrhea at presentation | Number of people in subject's household >17 years old | School attendance |
| Abnormal HR at presentation | Abnormal RR at presentation | Oral rehydration before presentation | Antibiotics taken before presentation |
| Number of vomiting episodes in last 24 hours prior to presentation | Other antibiotics | Antidiarrheal medication before presentation | Swimming in past month |
| Seizures | Contact with animal/pet in last month | Out of state travel in past month | Care prior to enrollment visit |
| Headache | Sick contact with GI illness in last month | Abnormal temperature at presentation | Member of household sought medical care for GI illness in past month |
| Constipation | Member of household sought medical care for GI illness in past month | Rotavax | Diarrhea had “other” description |

**Footnote:** Variables were selected based on availability at the time of emergency department presentation and potential clinical relevance for predicting viral-only etiology. Only predictors that could be obtain from caregiver history, physical examination, or routinely collected clinical information were considered for model development. ED; Emergency department.

**Supplementary Table 3**: Variable available to collect in emergency department (ED) and available in external validation dataset

| Variable considered in analysis | | | |
| --- | --- | --- | --- |
| Subject Age | Vomiting/nausea | International travel in past month | Immunosuppressive condition |
| Fever (at presentation) | Fatigue/weakness | Duration of vomiting at presentation | Duration of diarrhea at presentation |
| Diarrhea bloody | Number of vomiting episodes in last 24 hours prior to presentation | Excessive crying/fussiness | School attendance |
| Out of state travel in past month | Number of stools in 24 hours prior to presentation | Contact with animal/pet in last month | Antibiotics taken before presentation |
| season | Member of subject's household sought medical care for their GI illness | Number of people in the subject's household less than 5 yrs. old | Number of people in the subject's household between 5 and 17 yrs. Old |
| Member of subject's household sought medical care for their GI illness | Care prior to enrollment visit | Subject has received rotavirus vaccine in past month | Temperature (14-day average) |
| CO (14-day average) |  |  |  |

**Footnote:** Table lists candidate predictors that were available at the time of emergency department presentation and were also collected in both the derivation (IMPACT) and external validation (APPETITE) DATASET. Only variables available in both were considered for model development and external validation.

**Supplementary Figure 1:** Study Flow Chart
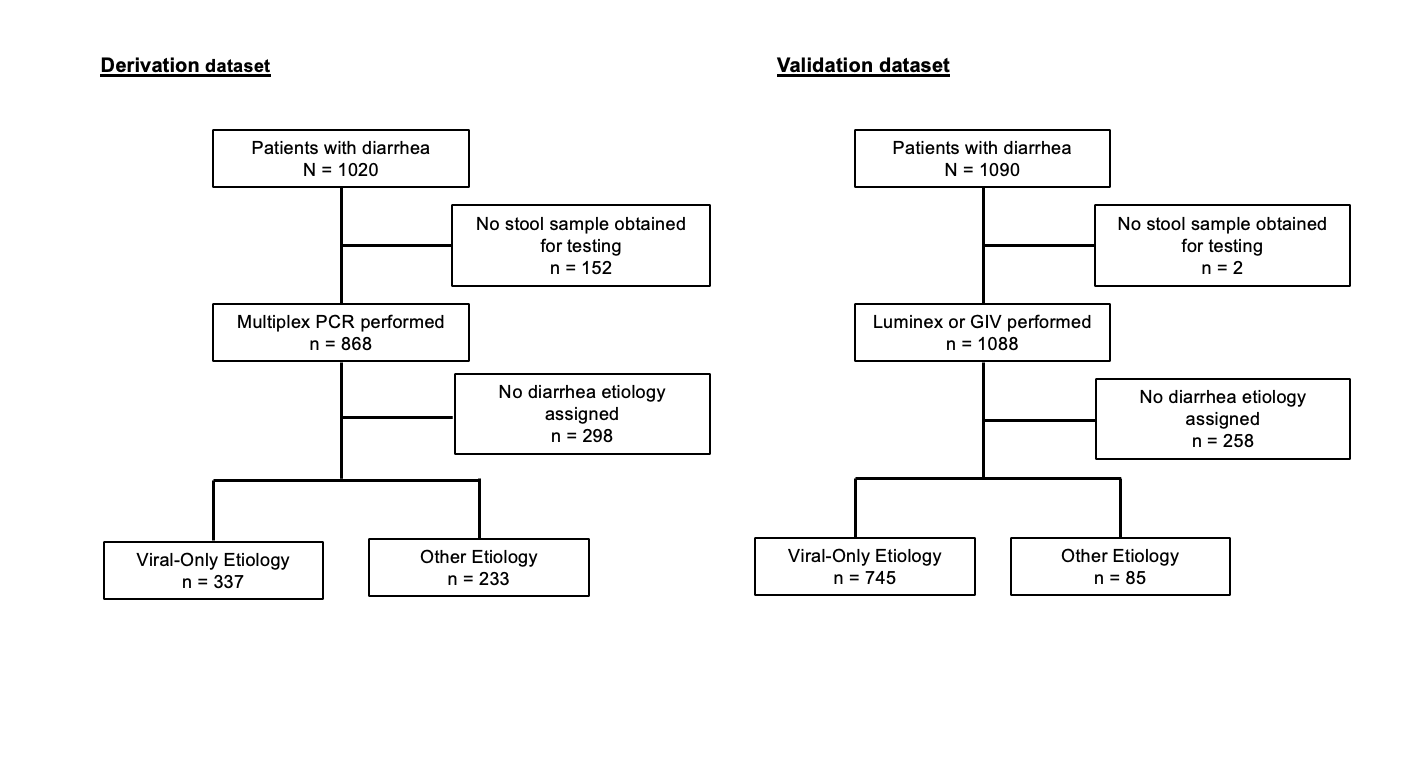

**Footnote:** For the derivation study Multiplex PCR testing was performed for pathogen detection. In validation study Luminex was performed for pathogen detection. Sapovirus and Astrovirus were identified using Gastrointestinal Viral (GIV) panel, as targets were not available in the Luminex multiplex PCR platform. Percentages are calculated using the total children with diagnostic testing performed as the denominator. Pathogens are not mutually exclusive.

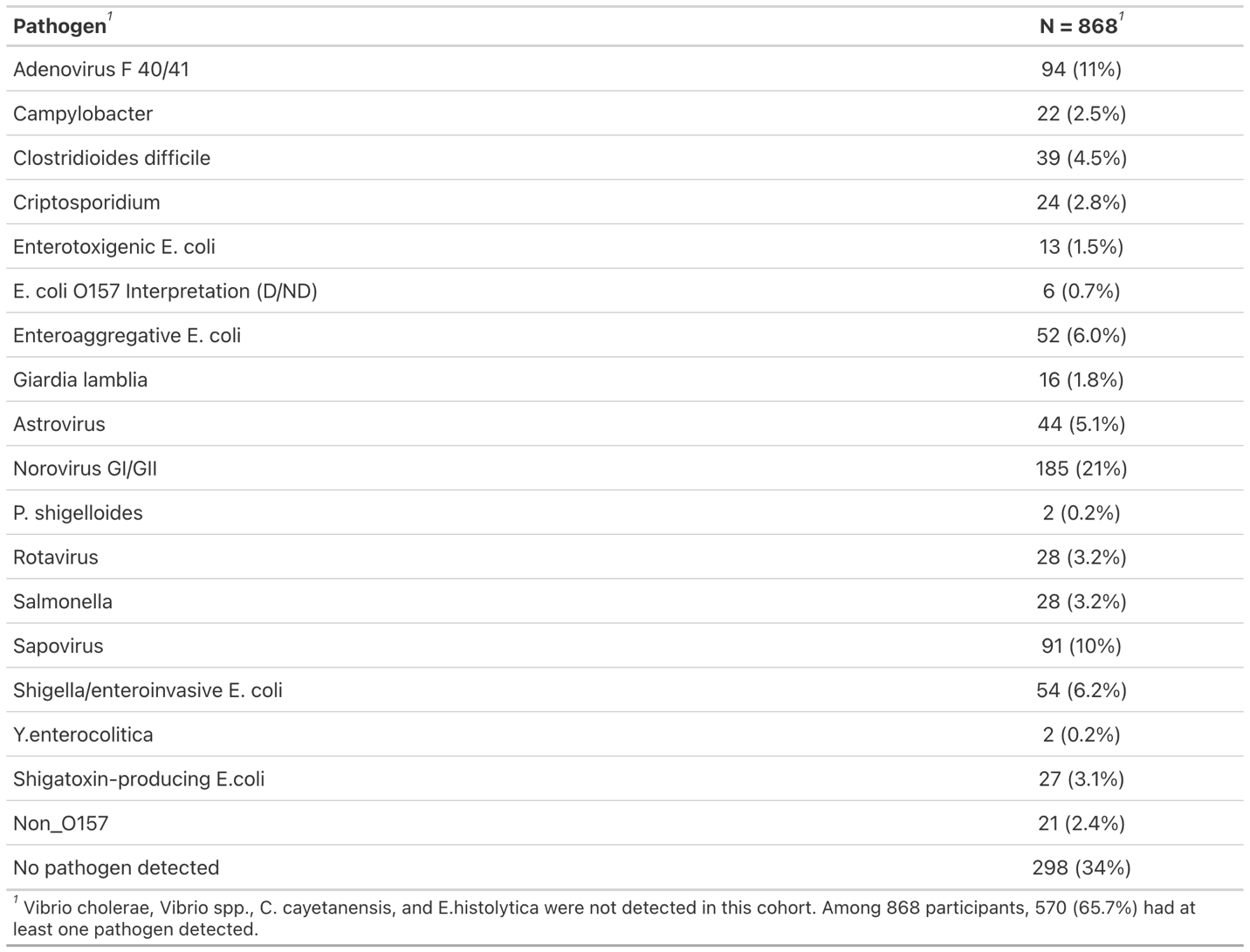
 **Supplementary Table 4:** Patients with diarrhea and PCR-performed in the IMPACT study

**Footnote:** This table summarizes the characteristics of children in the IMPACT cohort who presented with diarrhea and underwent multiplex polymerase chain reaction (PCR) testing for enteric pathogens. Only children with available PCR results were included in this subset. IMPACT, Implementation of Molecular Diagnostics for Pediatric Acute Gastroenteritis; PCR, polymerase chain reaction.

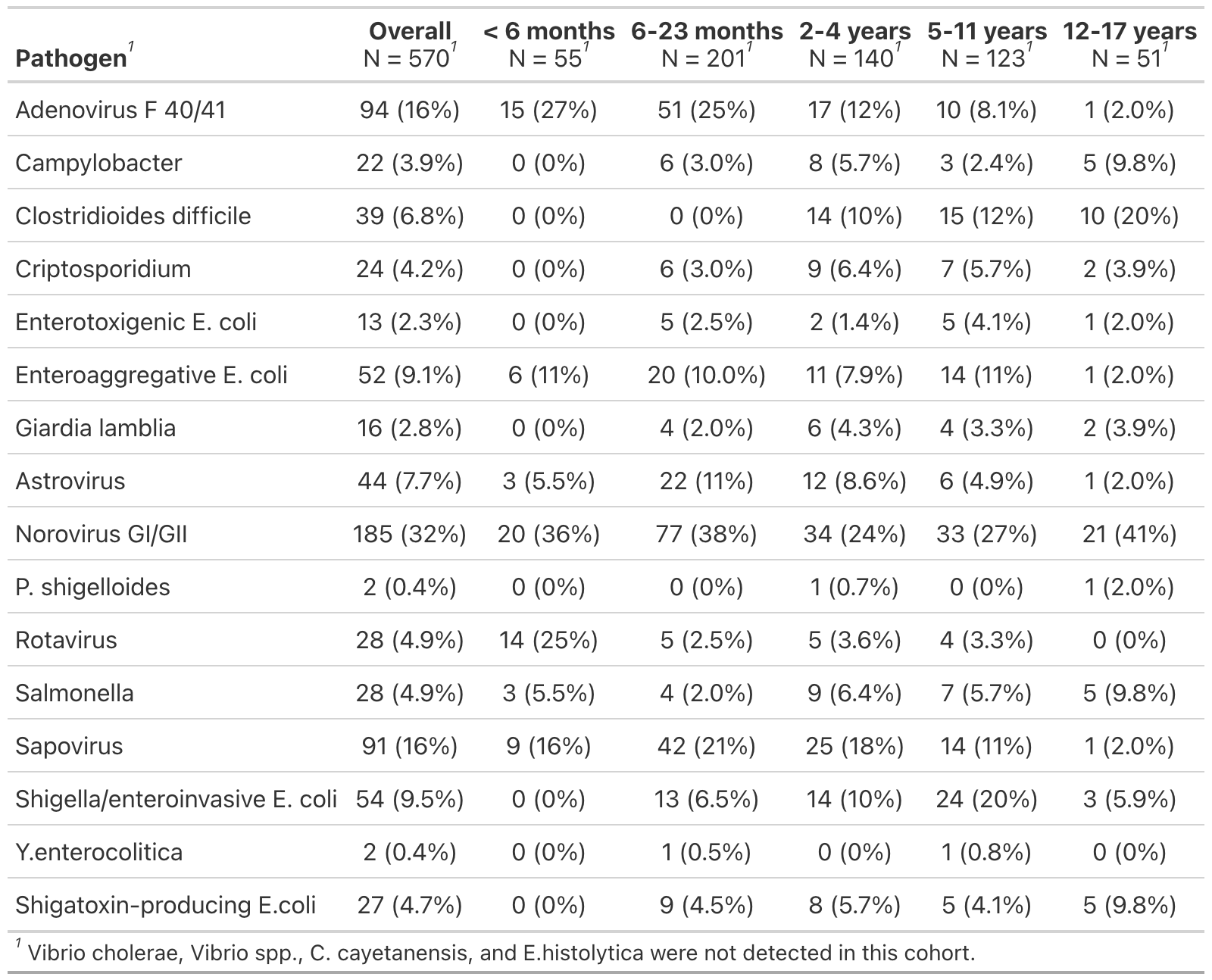
 **Supplementary Table 5:** Age-stratified distribution of PCR-detected pathogens among patients in the IMPACT study

**Footnote:** This table presents the distribution of enteric pathogens detected by multiplex polymerase chain reaction (PCR) across age groups in the IMPACT cohort. Multiple pathogens could be detected in the same patient; Therefore, percentage may sum to more than 100%. Age groups were defined as <6 months, 6-23 months, 2-4 years, 5-11 years, and 12-17 years. IMPACT, Implementation of Molecular Diagnostic for Pediatric Acute Gastroenteritis; PCR, polymerase chain reaction.

**Supplementary Table 6:** Patient distribution of diarrhea etiology among patients with PCR-performed (IMPACT and APPETITE datasets)

| **IMPACT & APPETITE**  **Dataset** | | | |
| --- | --- | --- | --- |
| **Pathogen Category** | **IMPACT**  **(n = 868)** | **APPETITE**  **(n = 1,088)** | **Final Classification** |
| Virus | 337 (38.82%) | 745 (68.4%) | Viral |
| Bacteria | 138 (15.90%) | 57 (5.2%) | Other |
| Bacteria + Virus | 56 (6.45%) | 22 (2.02%) | Other |
| Protozoa | 22 (2.53%) | 2 (0.18%) | Other |
| Protozoa + Bacteria | 8 (0.92%) | _ | Other |
| Protozoa + Virus | 7 (0.81%) | 4 (0.36%) | Other |
| Protozoa + Bacteria + Virus | 2 (0.23%) | _ | Other |
| Undetected/None | 298 (34.33%) | 258(23.7%) | Undetected |

**Footnote:** Tale summarizes the distribution of laboratory-confirmed diarrhea etiologies among children with available polymerase chain reaction (PCR), Luminex xTAG Gastrointestinal Pathogen Panel, and in-house viral reverse transcription polymerase chain (RT-PCR) panel. IMPACT, Implementation of Molecular Diagnostic for Pediatric Acute Gastroenteritis; APPETITE, Alberta Provincial Pediatric EnTeric Infections Team.

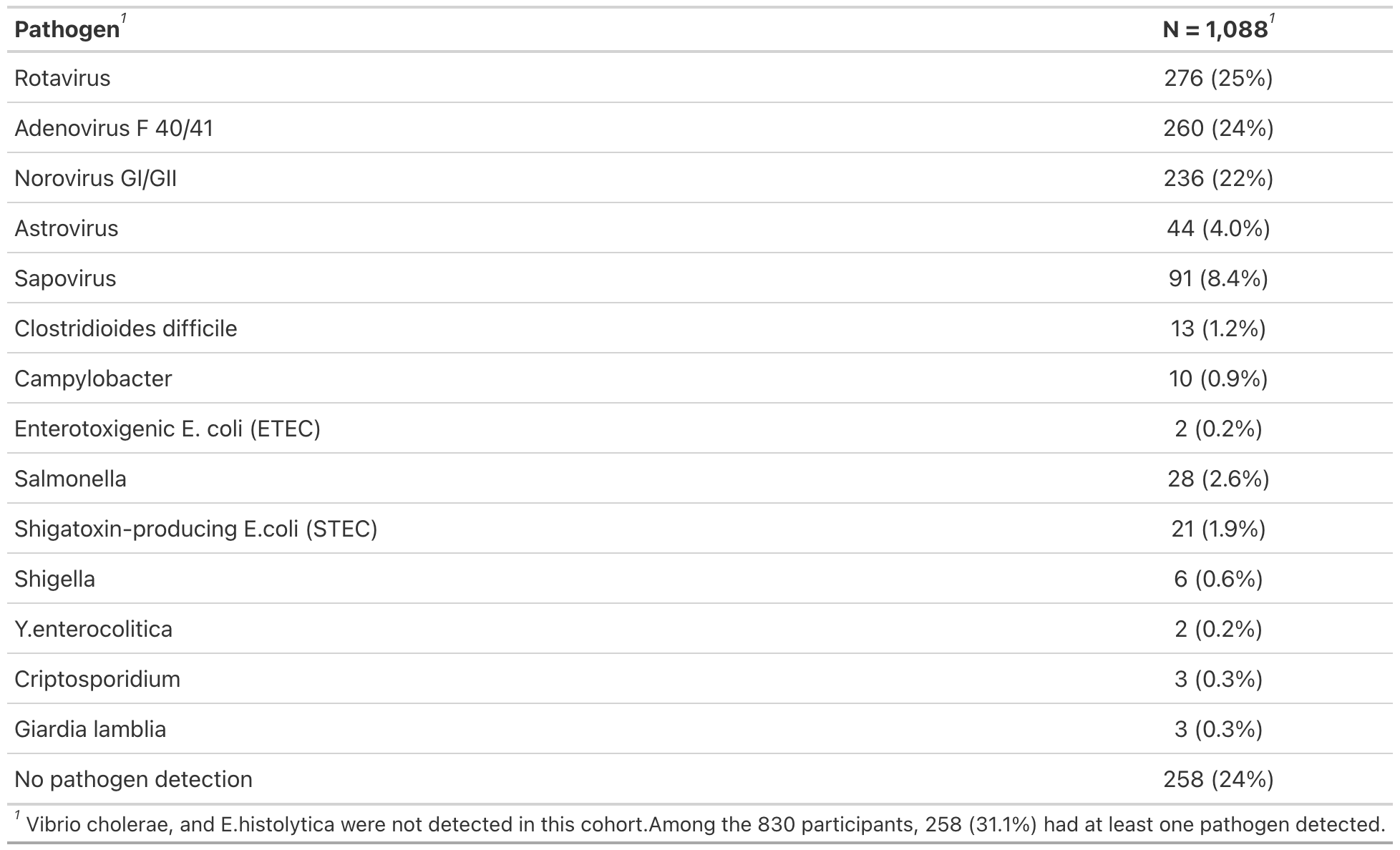
 **Supplementary Table 7:** Patients with diarrhea and PCR-performed in the APPETITE

**Footnote:** Table summarizes the characteristics of children in the APPETITE cohort who presented with diarrhea and underwent enteric pathogen testing, including Luminex xTAG Gastrointestinal Pathogen Panel, and in-house viral reverse transcription polymerase chain (RT-PCR) panel testing. Only children with available pathogen detection were included in the analysis. APPETITE, Alberta Provincial Pediatric Enteric Infection Team.

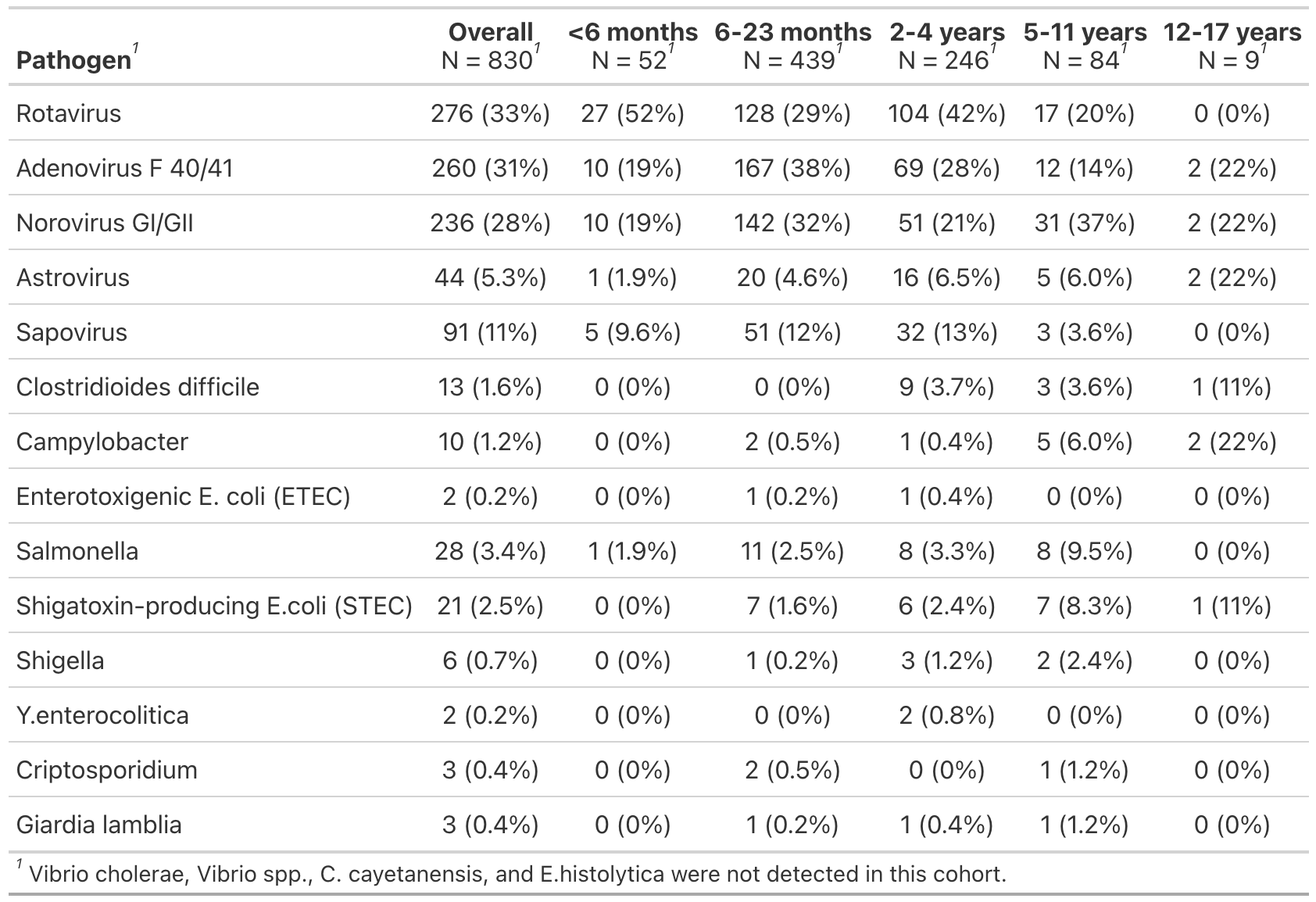
**Supplementary Table 8:** Age-stratified distribution of PCR-detected pathogens among patients in the APPETITE study

**Footnote:** This table presents the distribution of enteric pathogens detected by Luminex xTAG Gastrointestinal Pathogen Panel, and in-house viral reverse transcription polymerase chain (RT-PCR) panel testing across age groups in the APPETITE cohort. APPETITE, Alberta Provincial Pediatric Enteric Infection Team.

**Supplementary Figure 2:** Partial dependency plots for the top eleven predictors of viral etiology
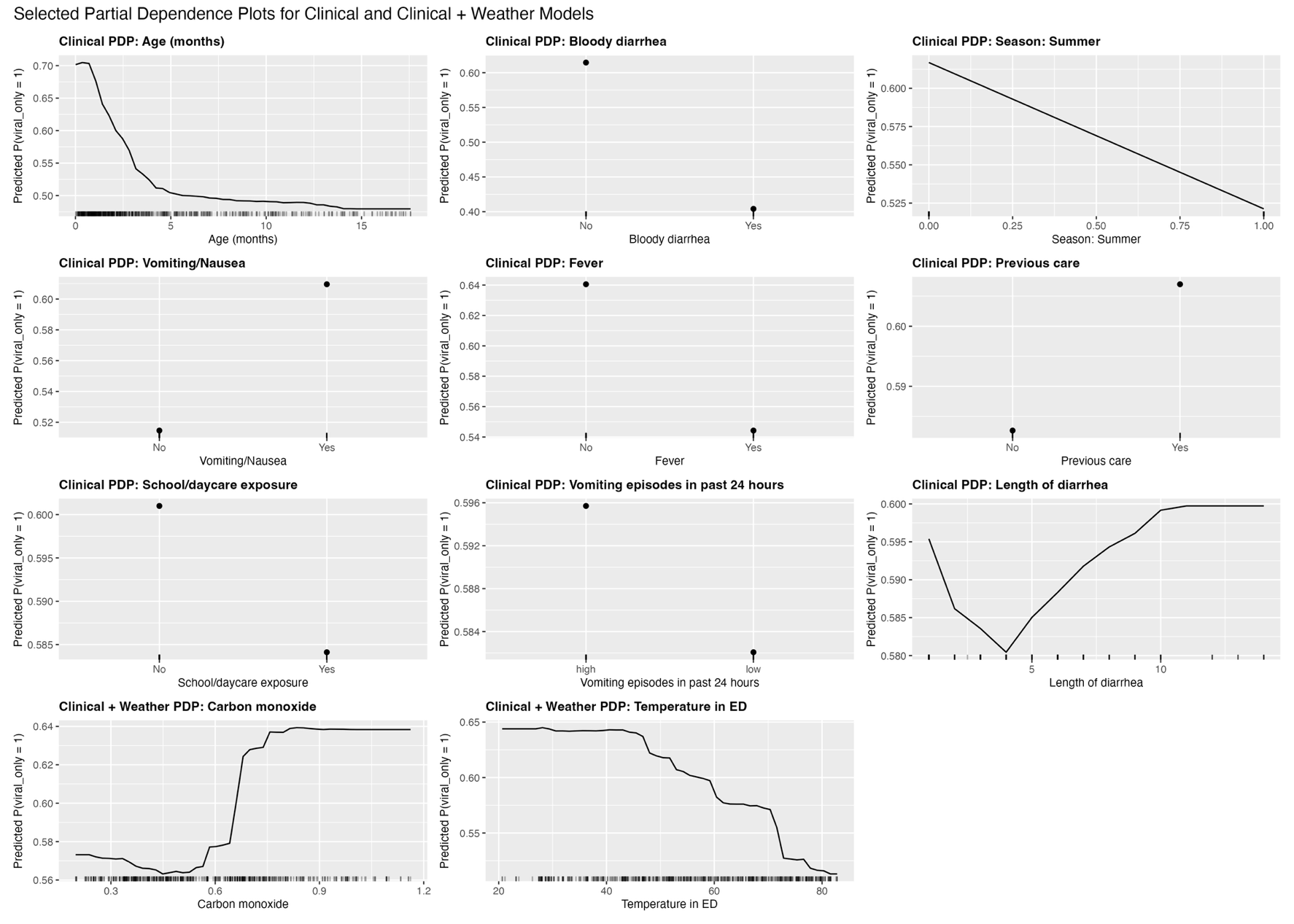

**Footnote:** Partial dependency plots show the average effect of each predictor on the predicted probability of viral-only etiology while holding all other predictors at their observed values. Plots were generated using the final random forest model. Viral etiology was defined as detection of one or more viral pathogens in the absence of bacterial or protozoal pathogens. Continuous predictors are displayed on their original scale, and categorical predictors are shown by level.

**Supplement Figure 3:** Ranking of clinical predictors based on AUC-based conditional permutation importance

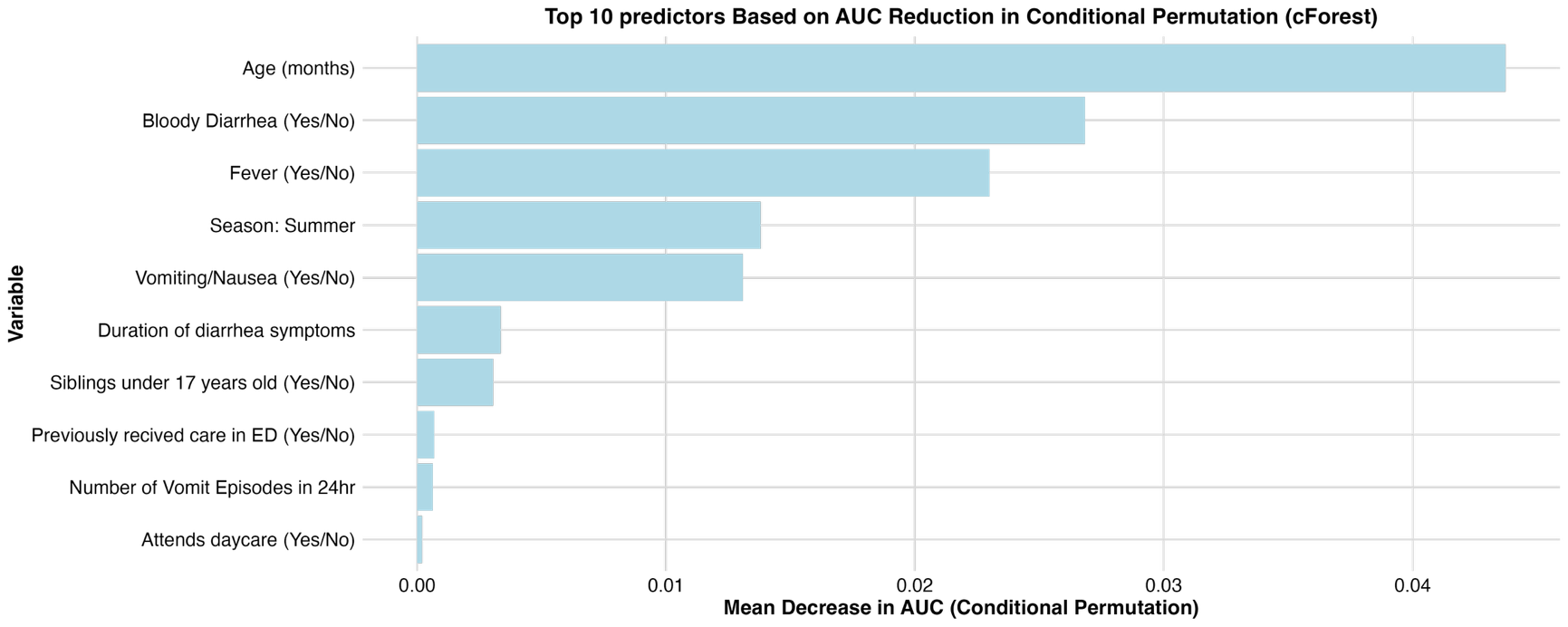

**Footnote:** Predictors were ranked according to the decrease in area under the receiver operating characteristic curve (AUC) after permutation of each variable using conditional inference random forest. Higher importance values indicate greater contribution to model discrimination.

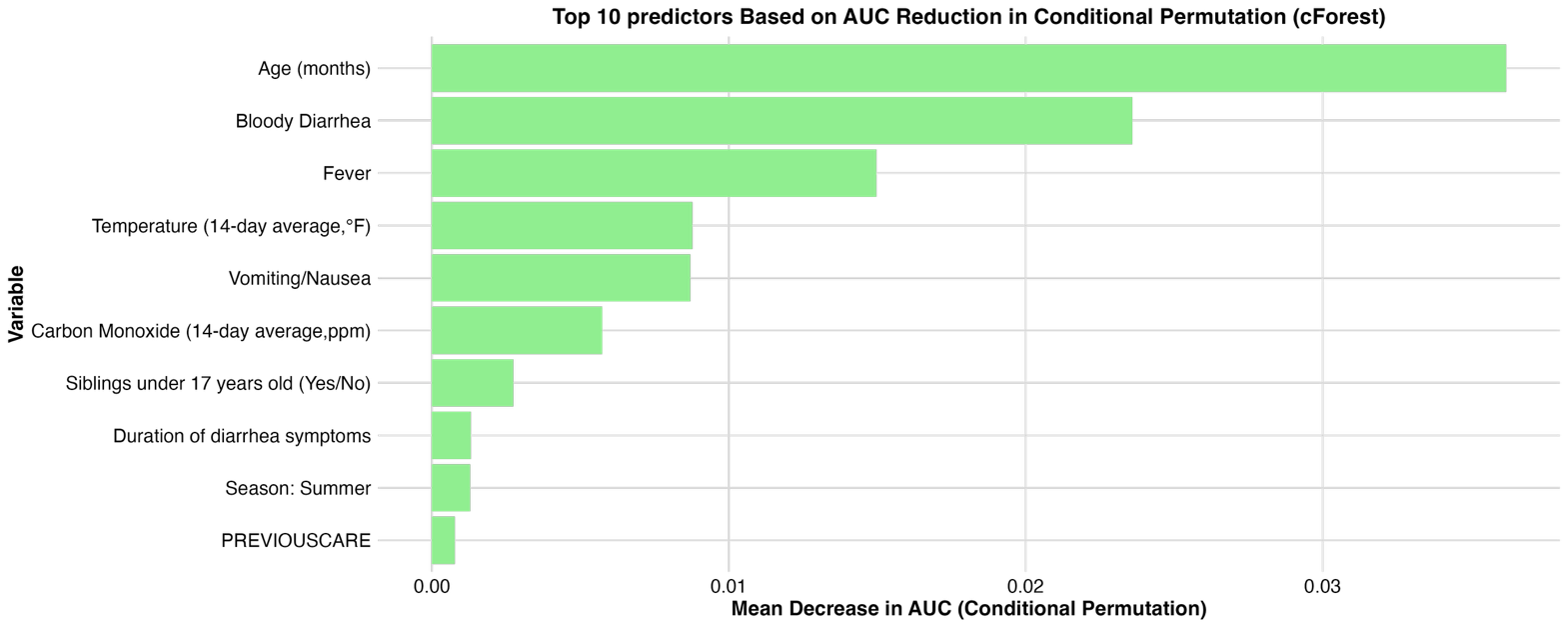
**Supplement Figure 4:** Ranking of clinical + Environmental predictors based on AUC-based conditional permutation importance

**Footnote:** Predictors were ranked according to the decrease in area under the receiver operating characteristic curve (AUC) after permutation of each variable using conditional inference random forest. Higher importance values indicate greater contribution to model discrimination.

**Supplementary Table 9:** External validation performance of clinical-only and clinical + environmental LR and RF models with varying predictor sets in the APPETITE dataset

| Number of predictors | AUC | Sensitivity | Specificity | PPV | NPV |
| --- | --- | --- | --- | --- | --- |
| Clinical-only model |  |  |  |  |  |
| Logistic regression |  |  |  |  |  |
| 3 | 0.767 | 0.915 | 0.482 | 0.939 | 0.394 |
| 5 | 0.811 | 0.911 | 0.552 | 0.947 | 0.415 |
| 10 | 0.799 | 0.849 | 0.600 | 0.949 | 0.312 |
| Random forest classification |  |  |  |  |  |
| 3 | 0.764 | 0.908 | 0.517 | 0.942 | 0.392 |
| 5 | 0.804 | 0.895 | 0.541 | 0.944 | 0.370 |
| 10 | 0.768 | 0.820 | 0.529 | 0.938 | 0.251 |
| Clinical + environmental model |  |  |  |  |  |
| Logistic regression |  |  |  |  |  |
| 3 | 0.767 | 0.915 | 0.482 | 0.939 | 0.394 |
| 5 | 0.790 | 0.940 | 0.400 | 0.932 | 0.435 |
| 10 | 0.783 | 0.849 | 0.600 | 0.949 | 0.312 |
| Random forest classification |  |  |  |  |  |
| 3 | 0.764 | 0.908 | 0.517 | 0.942 | 0.392 |
| 5 | 0.793 | 0.918 | 0.458 | 0.936 | 0.390 |
| 10 | 0.767 | 0.889 | 0.482 | 0.937 | 0.333 |

**Footnote:** PPV = positive predictive value; NPV = negative predicted values. Logistic regression (LR) and random forest (RF) models were developed using the IMPACT derivation cohort and externally validated in the APPETITE cohort (n=830). Predictor sets included the top 3, 5, and 10 variables selected by variables importance ranking. Performance measures included AUC, sensitivity, specificity, PPV, and NPV.

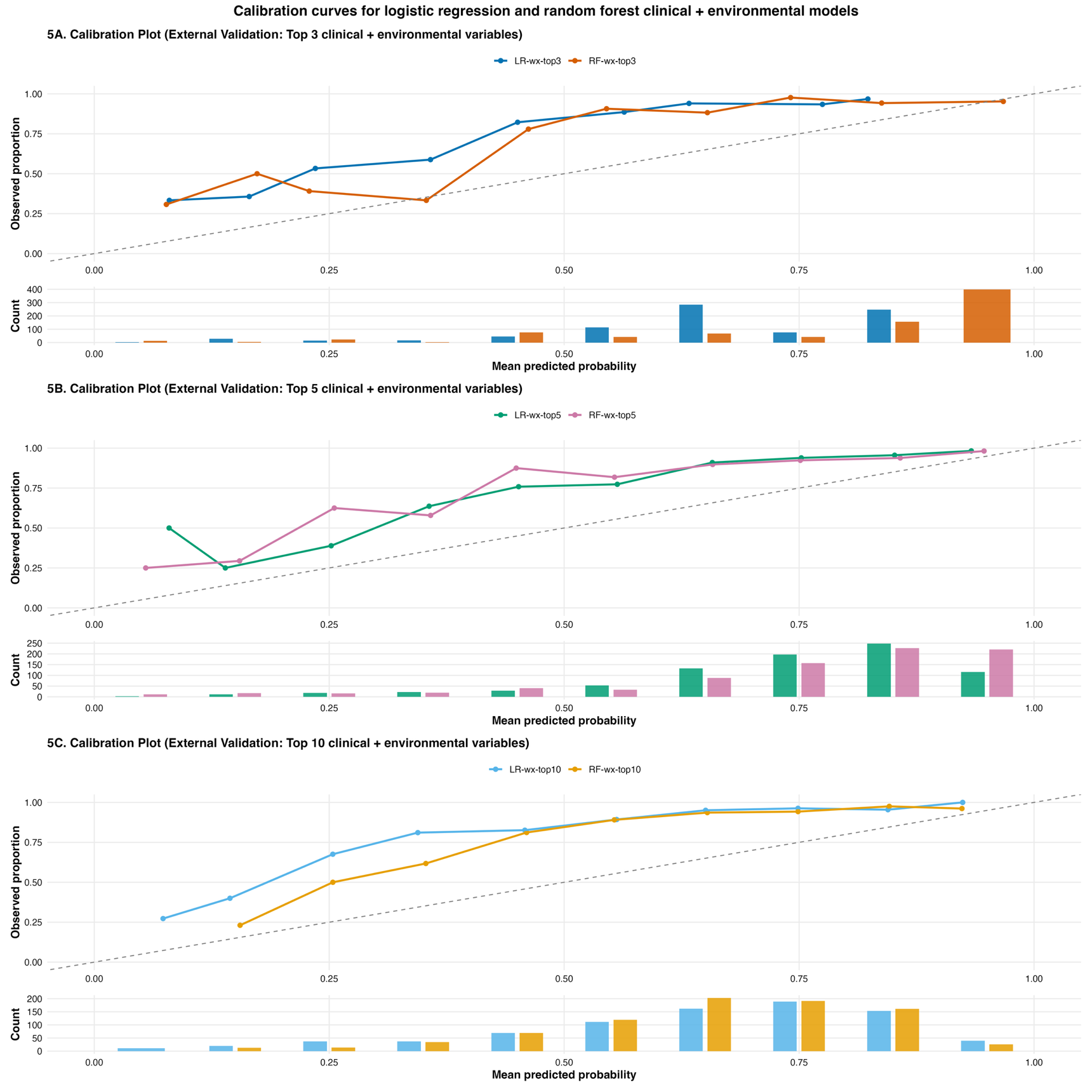
**Supplemental Figure 5:** External validation calibration plots for clinical + weather prediction models using 3, 5, and 10 predictors (APPETITE dataset).

**Footnote:** Calibration plots compare predicted probabilities of viral-only etiology (x-axis) with the observed proportion of viral-only cases (y-axis). Panels show models including models of clinical plus environmental variables with top 3, 5, and 10 predictors. The 45-degree reference line represents perfect calibration, where predicted probabilities correspond exactly to observed outcome frequencies. Deviations above line indicate underestimation, whereas deviations below the line indicate overestimation of the probability of viral-only etiology.

**Supplementary Table 10:** Calibration performance (intercept and slope) of logistic regression and random forest models across varying numbers of predictors

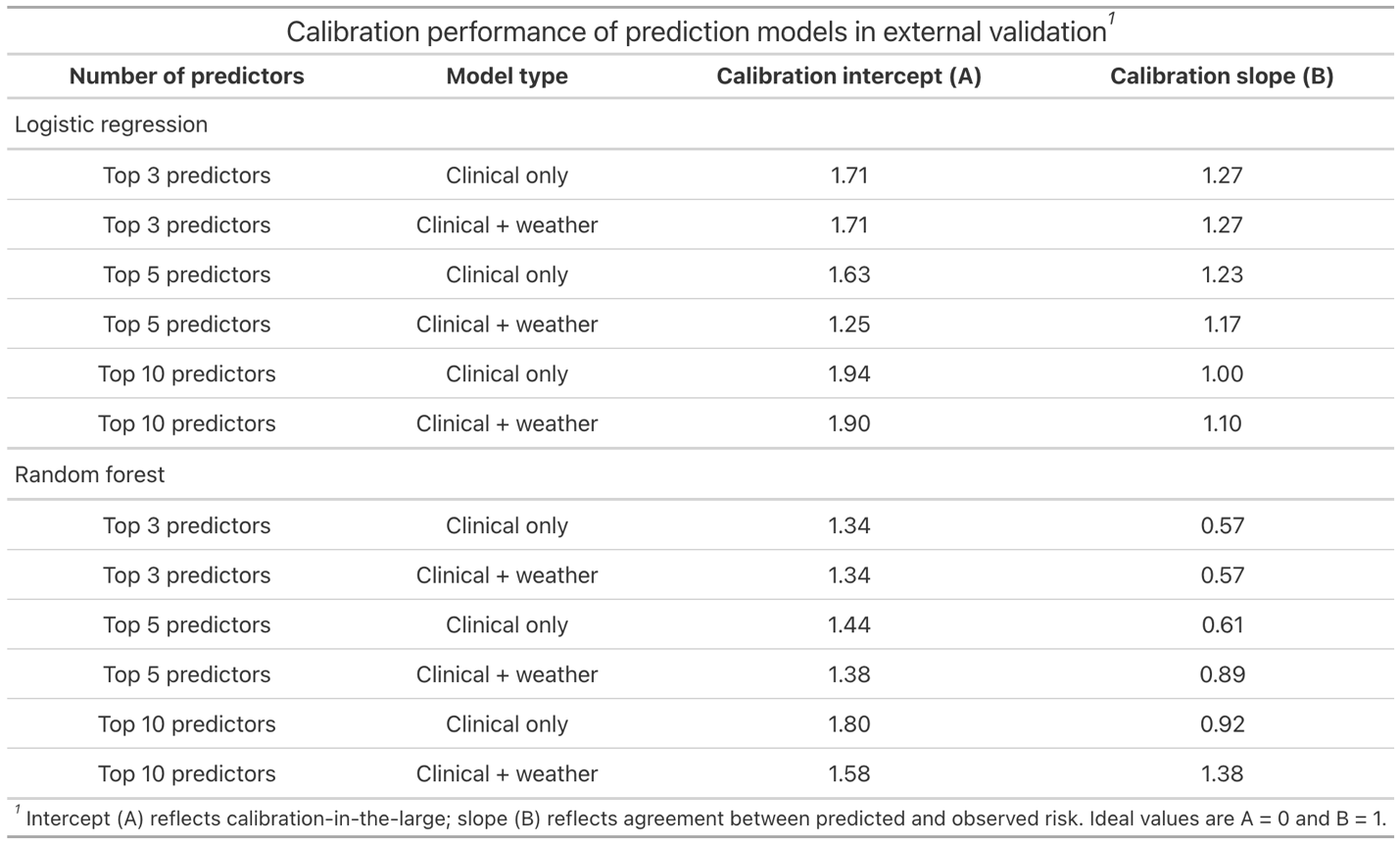

**Footnote:** Calibration performance was assessed using calibration intercept and slope. An intercept of 0 and slope of 1 indicate ideal calibration. Positive intercept values indicate underestimation, negative values indicate overestimation, and slope less than 1 suggest overfitting. Results are shown for logistic regression and random forest models across varying numbers of predictors.
